# XplainScar: Explainable Artificial Intelligence to Identify and Localize Left Ventricular Scar in Hypertrophic Cardiomyopathy from 12-lead Electrocardiogram

**DOI:** 10.1101/2024.05.22.24307764

**Authors:** Kasra Nezamabadi, Sanjay Sivalokanathan, Ji Won Lee, Talha Tanriverdi, Meiling Chen, Daiyin Lu, Jadyn Abraham, Neda Sardaripour, Pengyuan Li, Parvin Mousavi, M. Roselle Abraham

## Abstract

Left ventricular (LV) scar is a risk factor for sudden cardiac death and heart failure in hypertrophic cardiomyopathy (HCM). LV scar is frequent in HCM and evolves over time. Hence there is a need for LV scar detection and longitudinal monitoring. The current gold standard for LV scar detection is late gadolinium enhancement (LGE) on magnetic resonance imaging (MRI), which is limited by high cost and susceptibility to artifacts from implanted defibrillators. We introduce *XplainScar*, the first explainable machine learning method for LV scar detection and localization in HCM, using 12-lead electrocardiogram (ECG) data, which is not influenced by implanted devices. We use 500 patients from the JH-HCM Registry for model development, and 248 patients from the UCSF-HCM-Registry for validation. *XplainScar* combines unsupervised and self-supervised ECG representation learning, resulting in high precision (90%), sensitivity (95%), specificity (80%) and F1-score (90%) for scar detection in the basal, mid, and apical LV myocardium, with a processing time of <1 minute per 10 patients. Basal LV scar prediction by *XplainScar* is dominated by QRS features, and mid/apical LV scar by T wave features. *XplainScar* generalizes well to the held-out test UCSF data, with 88% precision, 90% sensitivity, 78% specificity, and F1-score of 89%. In summary, *XplainScar* demonstrates good performance for LV scar detection, and provides ECG signatures of basal, mid, and apical LV scar in HCM.

*XplainScar* is publicly available at https://github.com/KasraNezamabadi/XplainScar

## Introduction

Hypertrophic cardiomyopathy (HCM), the most common cardiac genetic disease and cause of sudden cardiac death in young individuals, is characterized by variable penetrance and phenotypic heterogeneity.^1–8^ The pathologic hallmarks of HCM are myocyte hypertrophy, myocyte disarray, cardiac fibrosis (scar), microvascular remodeling. But the location/extent of left ventricular hypertrophy (LVH) and scar vary, even among individuals from the same family, and evolve over time.^9,10^ Longitudinal monitoring of LV scar and LVH by magnetic resonance imaging (MRI) is recommended by the AHA/ACC guidelines [9] because high LV scar burden (>15% of LV mass) and severe LVH (wall thickness >3 cm) are risk factors for sudden cardiac death and heart failure.^9–13^ However, the high cost and limited availability of MRI worldwide, as well as susceptibility to artifacts from implanted defibrillators complicate longitudinal scar monitoring by MRI.^14^ Hence there is need for MRI-independent methods for LV scar detection in HCM.

The 12-lead electrocardiogram (ECG) is an ideal candidate for LV scar detection because it is widely available worldwide, relatively inexpensive, not influenced by defibrillator implantation, and reflects regional as well as global cardiac electrical activity that can be impacted by the presence of LV scar. Specifically, the QRS complex reflects myocardial depolarization, and the ST segment as well as T wave reflect repolarization; LV scar would be expected to primarily impact impulse propagation in the LV and hence the QRS complex. But, ischemic, dilated, and hypertrophic cardiomyopathies, which are associated with LV scar, can also have concomitant electrical remodeling, leading to changes in the ST segment and T wave in addition to QRS complex abnormalities.^15^ In the case of HCM, pathogenic variants in sarcomeric protein genes lead to myocyte hypertrophy, myocyte disarray, interstitial and/or replacement fibrosis, changes in ion channel/gap junction expression/function and/or coronary microvascular structure/function, which can influence myocardial depolarization and repolarization.^16–18^ This structural and/or electrical remodeling in HCM hearts leads to a variety of ECG abnormalities involving the QRS complex, ST segment and T wave in several or all leads, that evolve over time.^19–23^ But unlike myocardial infarction where LV scar is often transmural and in a vascular distribution, LV scar in HCM is often patchy, mid-myocardial and not in a vascular territory.^24^ This could explain why Q waves which are highly predictive of LV scar in ischemic cardiomyopathy, do not predict LV scar in HCM.^25^ Small studies in HCM patients suggest an association between QRS complex fragmentation, low QRS voltage, T wave inversion and strain pattern with LV scar.^26,27^ But specific methods to predict LV scar from 12-lead ECG data in an individual with HCM are lacking. We address this problem with a machine learning (ML)-based solution that learns from extensive HCM-ECG data to discern patient-specific ECG features indicative of LV scar. Our model *XplainScar*, extracts comprehensive features from routine 12-lead ECGs using an HCM ECG-specific segmentation algorithm, and incorporates unsupervised as well as self-supervised representation learning, to yield an explainable ML framework that effectively predicts the presence/absence of LV scar and reveals ECG markers associated with scar location, in < 1 minute per 10 patients.^28^

## Methods

### Ethic Approval

The HCM Registries are approved by the Institutional Review Boards (IRB) of the Johns Hopkins (JH) and the University of California San Francisco (UCSF) Hospitals. Informed consent was obtained for use of medical records for research. Protected patient information was excluded from datasets used for machine learning methods.

### **Method Overview** (see Supplemental data section for detailed methods)

Cardiac MRI images and 12-lead ECG from HCM patients in the JH-HCM-Registry (*n* = 500) and UCSF-HCM-Registry ( *n* = 248 ) who underwent deep clinical phenotyping were used for model development and validation, respectively (**Suppl. Table 1, Suppl. Figure 1**). The ground truth for LV scar detection/localization was obtained by analysis of MRI images for late gadolinium enhancement (LGE) within 1 year of ECG; patients with LGE at right ventricular (RV) insertion points were excluded (**Suppl. Figure 1**) because LGE at this location may not represent scar.^29^

The input to our machine learning model *XplainScar* (**Figure 1**) is 276 features representing ventricular depolarization and repolarization, extracted from a segmented 12-lead HCM ECG, and adjusted for LV mass index, age, sex (**Suppl. Figure 2, Suppl. Table 2**).^28^ We perform unsupervised ECG clustering where HCM patients with similar ECG features are recursively partitioned into the same group, and groups where the LV scar group dominates the NoScar group and vice versa are merged until all patients are assigned to a merged group (**Figure 1, Suppl. Table 3**). For each resulting group, we train a self-supervised neural network followed by a fully connected neural net to predict the presence of scar in the basal, mid, and apical LV (**Figure 1**). Shapley values (**Suppl. Figure 3**) are used to explain LV scar prediction and gain insights into ECG signatures of LV scar in HCM.^30,31^ We evaluate model performance by computing specificity, sensitivity, positive predictive value, F1-score and 95% confidence interval for accuracy (**Table 1**).

**Figure 1.**
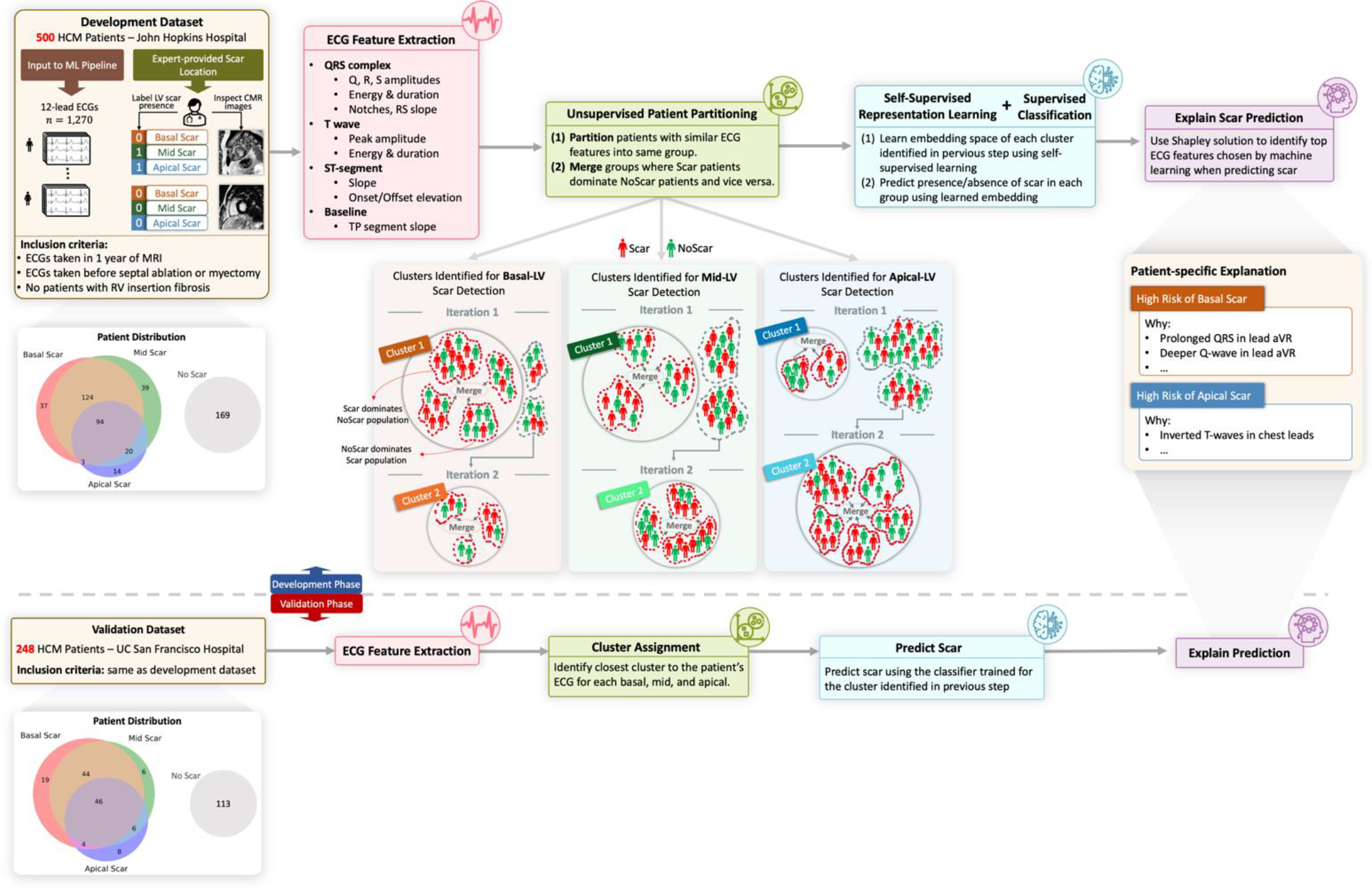
Overview of *XplainScar*. This method was developed using the JH-HCM dataset (n=500), and validated using the UCSF-HCM dataset (n=248), after excluding HCM patients with LGE at RV insertion sites. In each patient, the LV was divided into basal, mid, apical regions for scar (LV-LGE) detection. P, Q, R, S, T waves in 12-lead ECGs were identified using a segmentation method tailored for handling HCM ECGs. ECG features such as duration, amplitude, slope, energy of QRS complexes and T waves, as well as ST, TP segments were extracted from each lead, and adjusted for LV mass index, age, sex, using multiple linear regression. Subsequently, patients were partitioned into groups based on similarity of their ECG features using unsupervised clustering. In each group, a self-supervised neural network followed by a fully connected neural net predicted presence of LV scar. The Shapley value approach was used to identify the top ECG features that participated in LV scar prediction in basal, mid, apical LV in each patient.

**Table 1.**
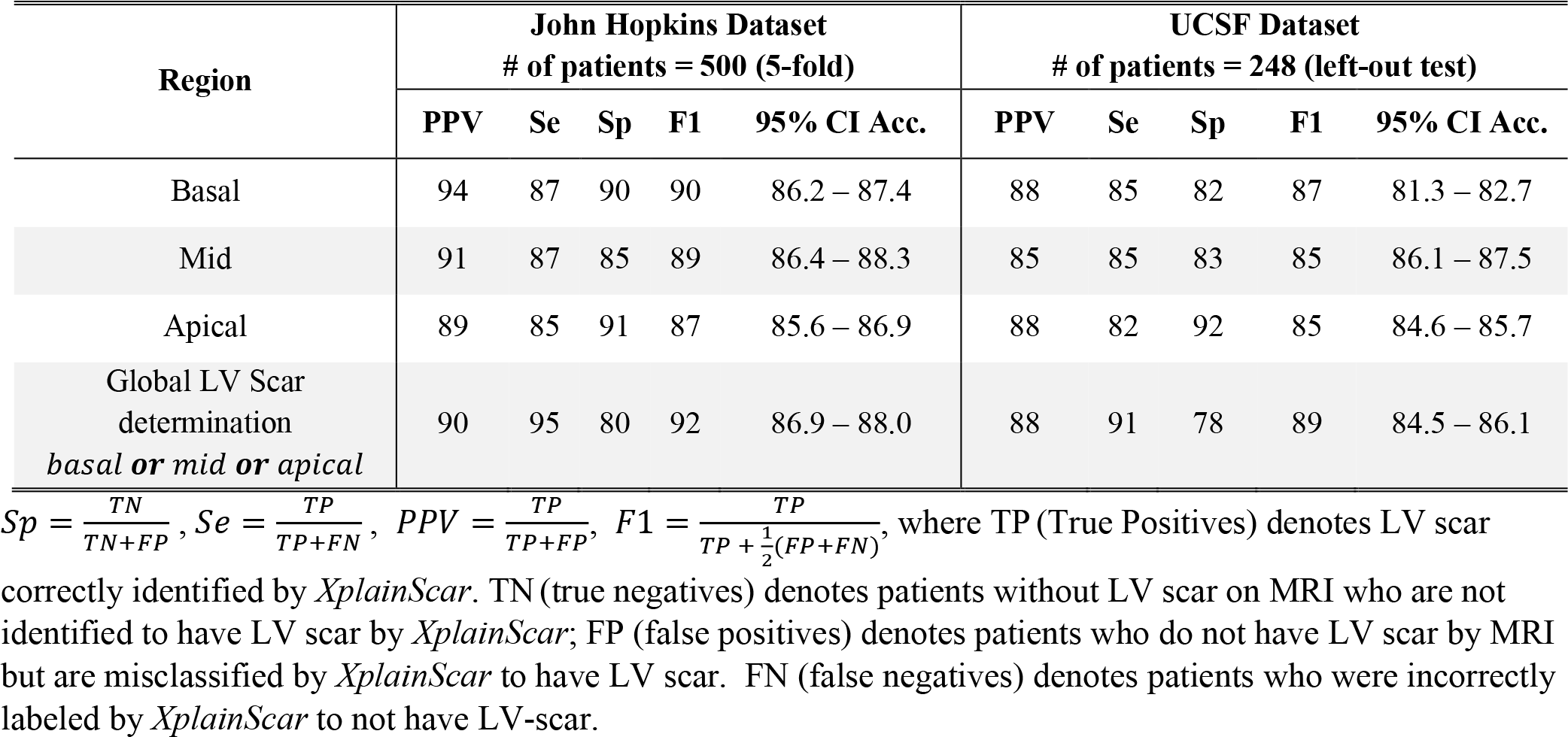
LV-scar detection performance by our method in terms of positive predictive value (PPV), sensitivity (Se), Specificity (Sp), F1 score (F1), and 95% confidence interval around accuracy.

## Results

### *XplainScar* can effectively detect LV scar using 12-lead ECG

We follow a 10-run, 5-fold cross-validation scheme for the JH dataset and exclusively test on the left-out UCSF dataset. Model performance for regional (base, mid, apex) and global LV scar detection is presented in **Table 1**. Our method achieved an F1 score of 0.92, sensitivity of 0.95, specificity of 0.8 over the JH dataset, and an F1 score of 0.89, sensitivity of 0.91, specificity of 0.78 over the left-out UCSF dataset. These results demonstrate the effectiveness of *XplainScar* in accurately identifying LV scar while highlighting its generalizability to unseen UCSF data. The small decrease in model performance when transitioning from the JH dataset to the UCSF set could result from factors such as differences in patient characteristics and data collection methods (MRI scanners) between the two centers. Notably, *XplainScar* requires <1 minute (wall clock) to predict LV scar in 10 HCM patients, using one 10-second 12-lead ECG per patient, on a personal computer (8 GB of RAM memory with no GPU). This swift processing time and robust performance on unseen UCSF data underscore the potential for practical deployment of *XplainScar* in real-world scenarios to assist with disease management in clinic.

### Ablation Experiments and Comparison with State-of-the-Art Methods

We performed ablation experiments (**Table 2**) to examine the importance of unsupervised ECG clustering and self-supervised neural network in determining model performance (F1 score). Elimination of the patient clustering step led to ∼50% reduction of the F1 score, and elimination of self-supervised representation decreased the F1 score by ∼10%. These results indicate that patient partitioning is a key step in our pipeline for LV scar detection.

**Table 2.**
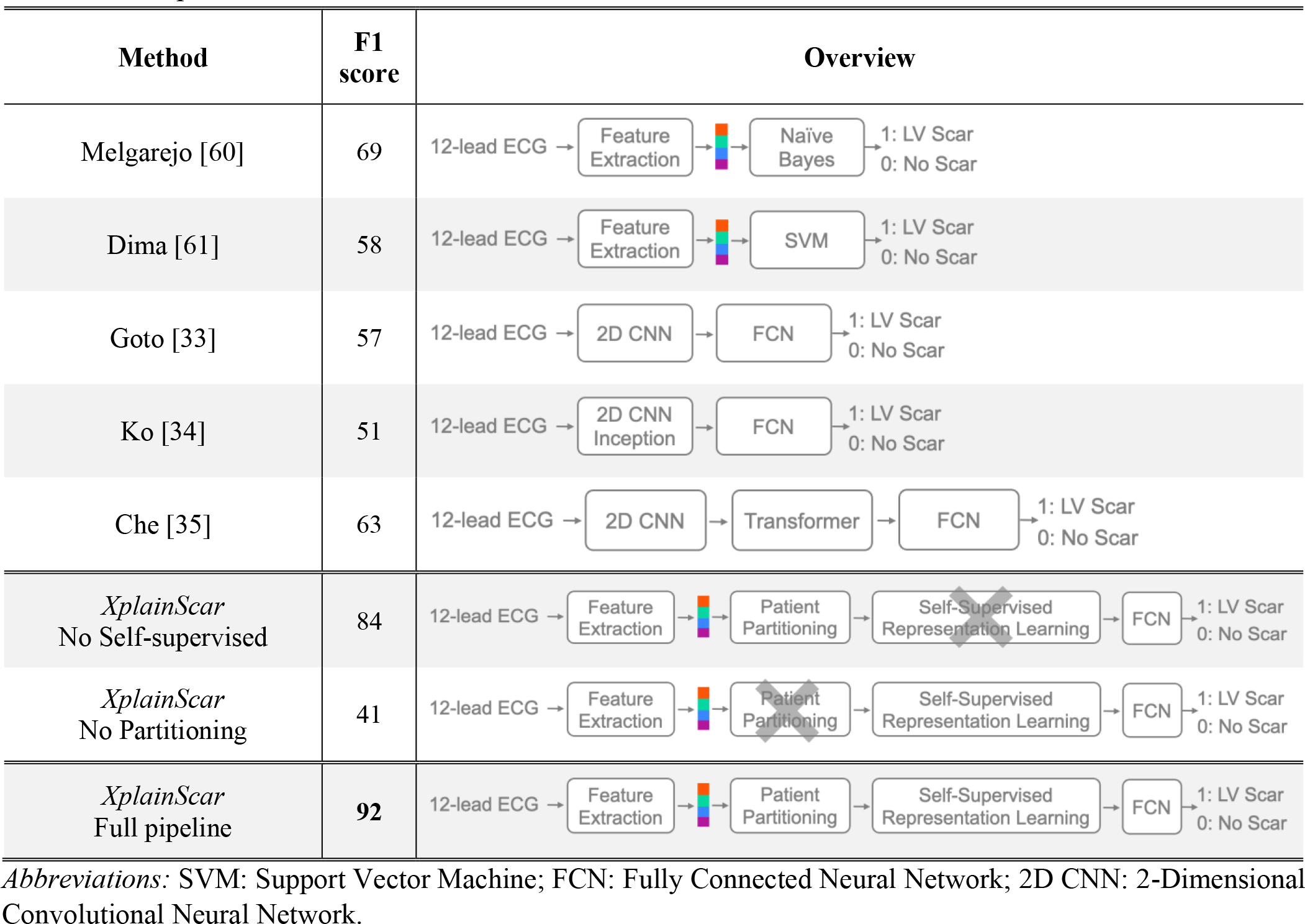
LV scar detection performance obtained by our method compared to five state-of-the-art methods and two ablation experiments over the JH dataset in terms of F1 score.

We did not find any deep learning approaches for ECG-based LV scar detection in HCM, so we leveraged state-of-the-art ECG-based deep neural networks proposed for HCM diagnosis and general arrhythmia detection for comparison (**Table 2**).^32–34^ The datasets used in these studies and the trained weights of the models are not publicly available; as such, we train the models over the JH dataset and test them following the same cross-validation scheme used by our method. To compensate for the relatively small size of the JH dataset, which can limit training of deep learning models, we synthetically generate 12-lead ECGs by employing the data augmentation method proposed by Gopal et al., where the 12-lead ECG is first transferred into 3D vectorcardiogram space using inverse Dowers transformation, randomly rotated, and then converted back to the ECG space.^35,36^ Furthermore, as the exact architecture of the compared deep models may not be optimal for LV scar detection (since they were originally proposed for disease diagnosis), we perform an extensive grid search to obtain the best hyperparameters, including the size and number of convolution kernels, attention heads, and layers. All the methods’ preprocessing steps, including ECG denoising and re-sampling, and feature extraction (for non-deep-learning methods) are implemented as the authors propose. Notably, *XplainScar* significantly outperforms all 5 methods (**Table 2**).

### XplainScar reveals ECG features of regional LV scar in HCM

We employ the Shapley additive explanation (SHAP) method to compute Shapley values for ECG features of LV scar.^31^ By denoting *XplainScar* as function *f* and the input ECG as *x* ∈ ℝ^*M*^ with *M* extracted features, we explain the prediction *f*(*x*) by assigning a Shapley value to each feature in *x* (**Suppl. Figure 3**). Each prediction is explained as a series of ECG features and Shapley value pairs where higher values show that the corresponding feature significantly impacts the prediction. We utilize the entire JH dataset as the training set for SHAP, which enables us to compute Shapley values for each prediction made on the UCSF dataset. **Table 3** shows the average Shapley value of each ECG component and corresponding ECG leads across all predictions. *XplainScar* identified several QRS-related features in leads I, V1, aVR as predictors of basal scar, and T wave related features in leads V4 – V6, for mid and apical LV scar detection.

**Table 3.**
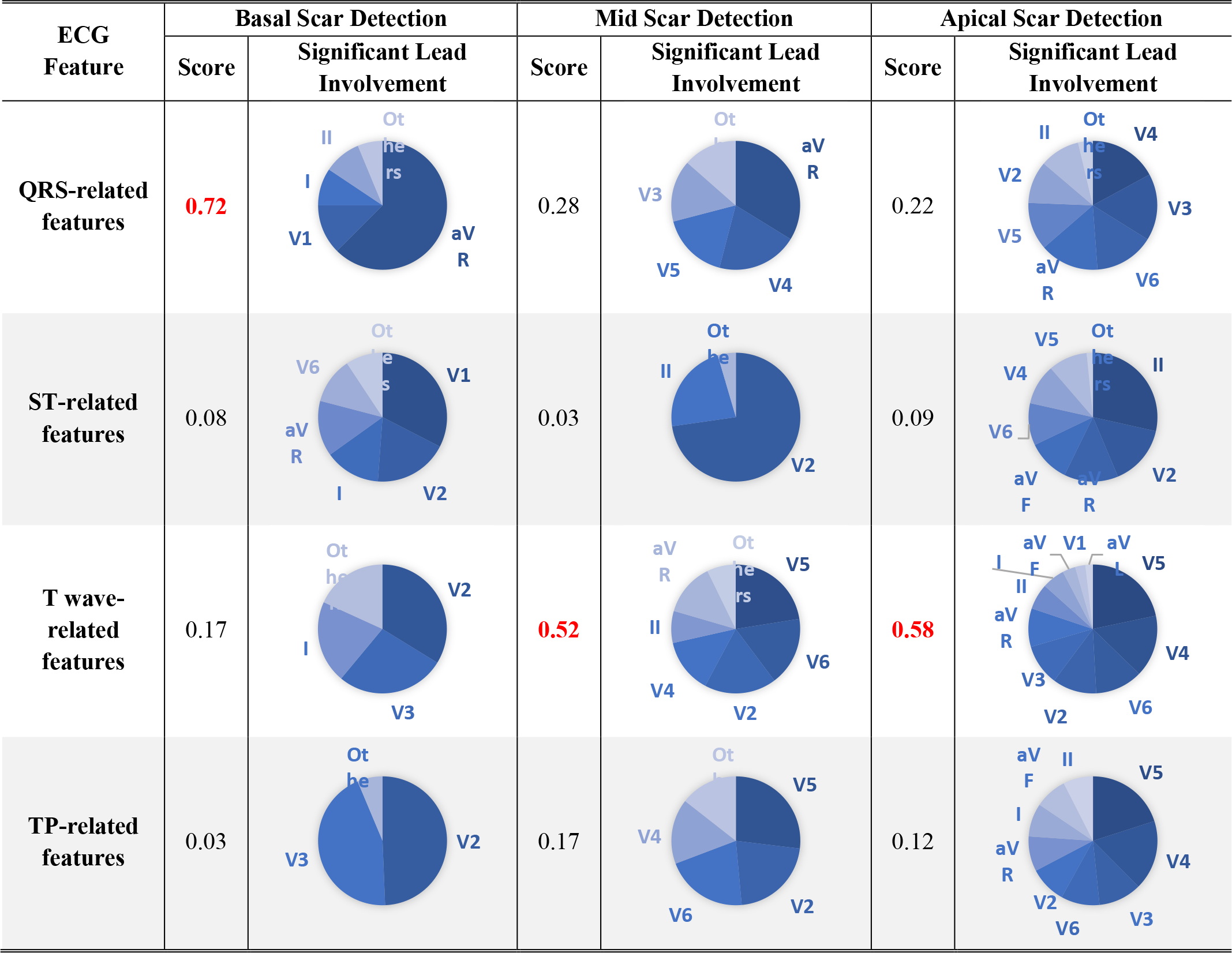
ECG Feature importance score calculated as the average Shapley value of each ECG component across all predictions in the UCSF dataset along with significant leads involvements.

| ECG Feature | Basal Scar Detection |  | Mid Scar Detection |  | Apical Scar Detection |  |
| --- | --- | --- | --- | --- | --- | --- |
|  | Score | Significant Lead Involvement | Score | Significant Lead Involvement | Score | Significant Lead Involvement |
| QRS-related features | **0.72** |  | 0.28 |  | 0.22 |  |
| ST-related features | 0.08 |  | 0.03 |  | 0.09 |  |
| T wave-related features | 0.17 |  | **0.52** |  | **0.58** |  |
| TP-related features | 0.03 |  | 0.17 |  | 0.12 |  |

To reduce the sensitivity of our analysis to large variations in Shapley values generated by SHAP for regional scar prediction, we count the number of predictions where the specific ECG feature appeared within the top 20% of Shapely values, and report those with the highest frequencies (instead of averaging Shapley values of a feature across predictions) (**Table 4**). The mean and standard deviation for each feature in the Scar group compared with the NoScar group, using Welch’s T-test, are presented in **Table 4**. The scatter plot (**Table 4)** further illustrates how the value of each feature impacts the model output, pushing it toward predicting Scar or NoScar. For example, the first row of **Table 4** shows that a deeper Q wave pushes *XplainScar* toward predicting basal scar, whereas absence of Q is associated with absence of basal scar (each dot in scatter plots corresponds to a single prediction).

**Table 4.**
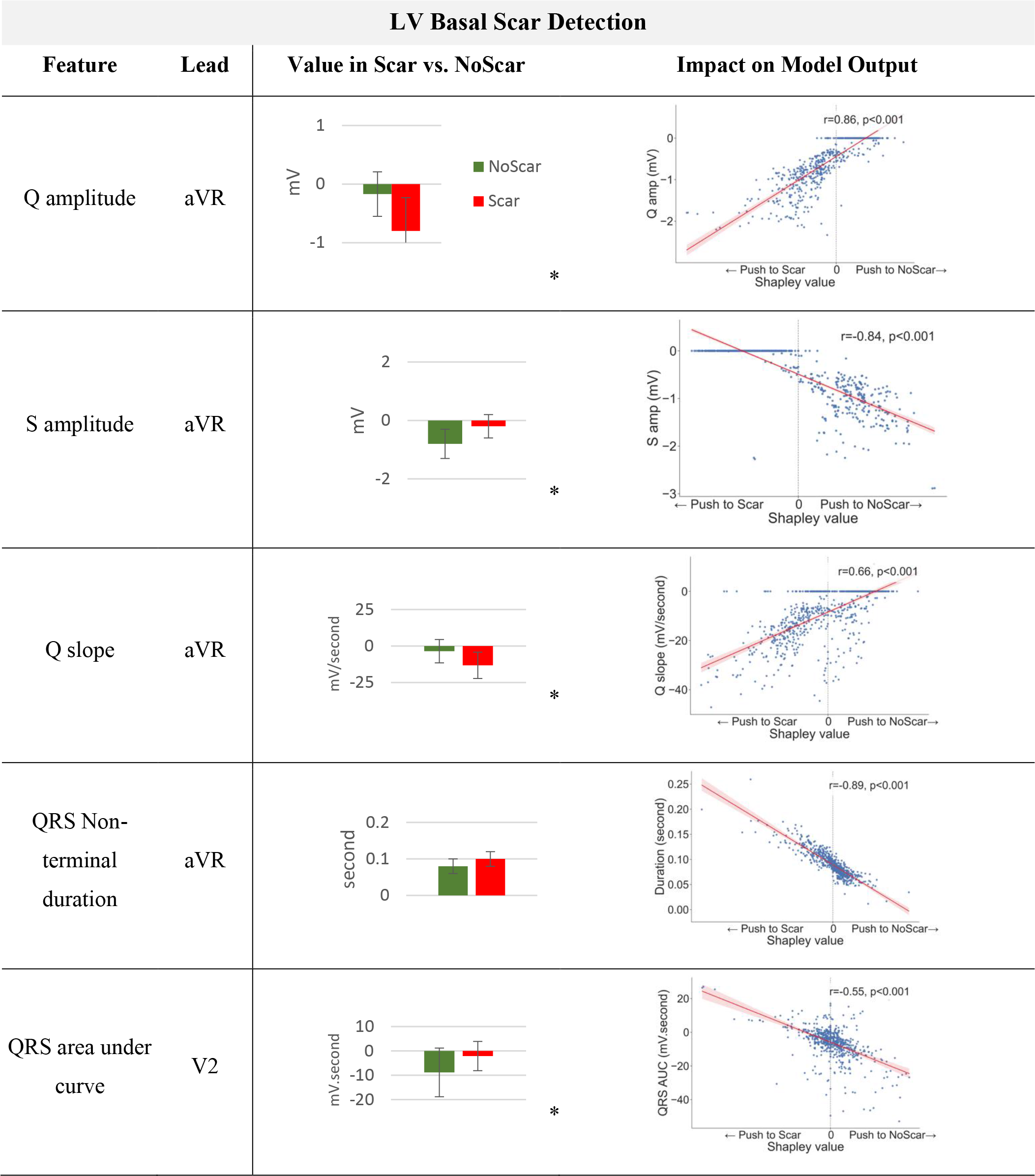
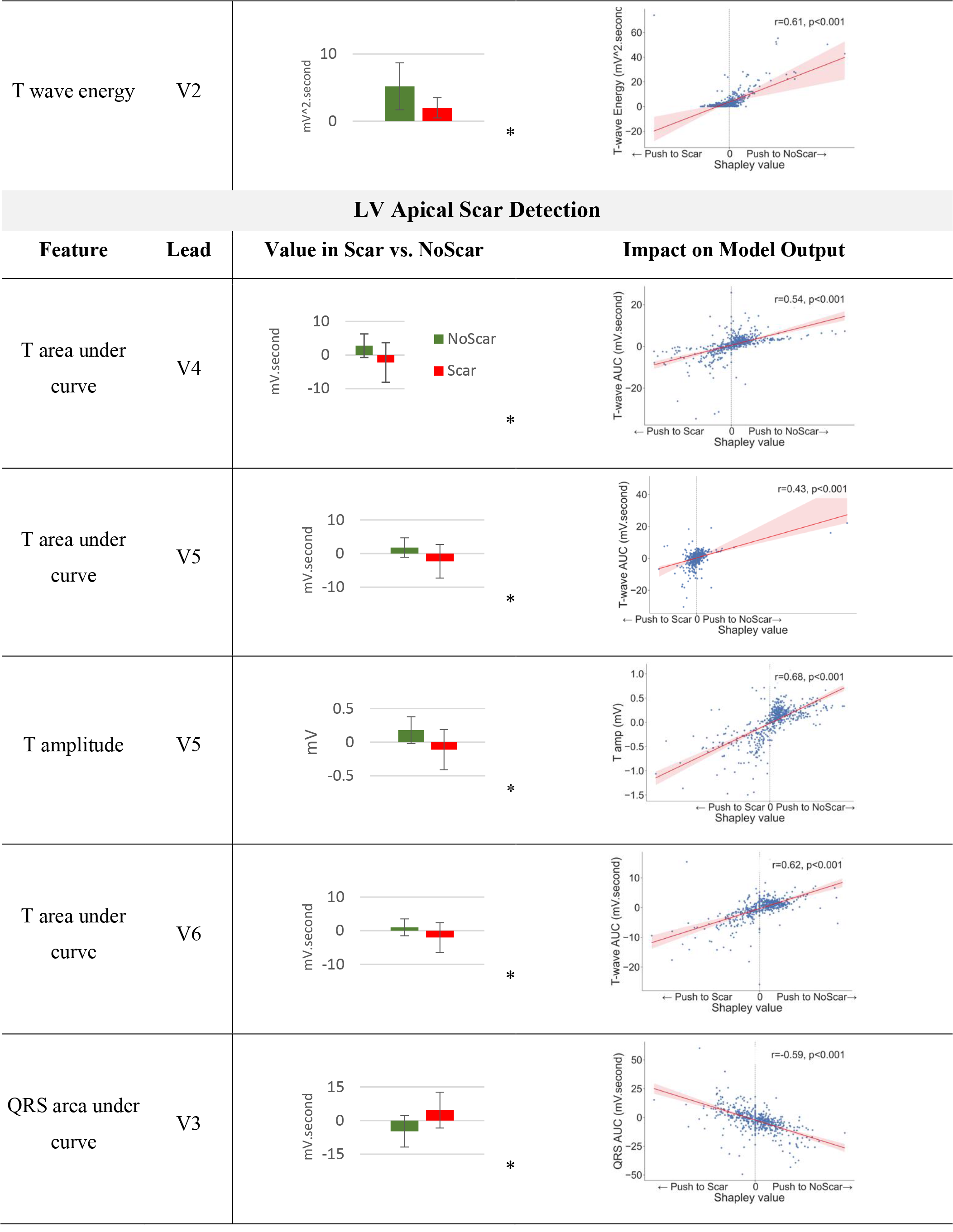
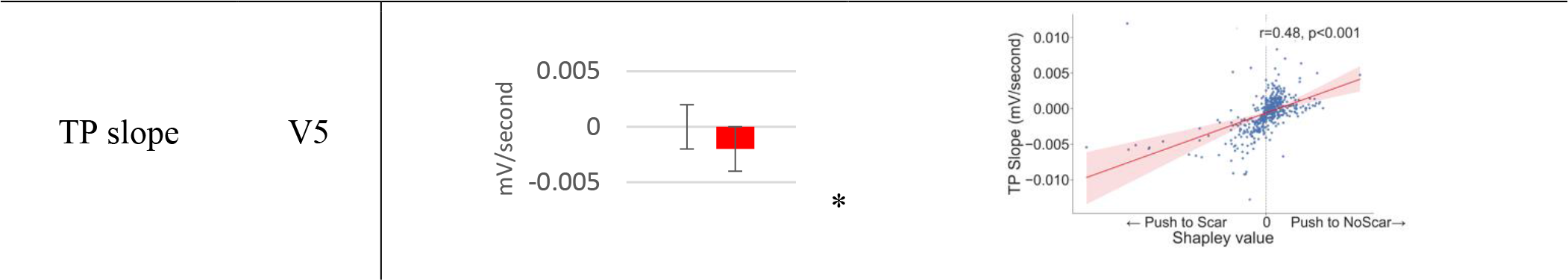
Top ECG features frequently selected by *XplainScar* when detecting basal and apical LV scar. Asterisk (*) shows statistically significant difference (p<0.05) between feature values in Scar vs. NoScar groups, obtained using Welch’s t-test. Each dot in scatter plots corresponds to a single prediction by *XplainScar*. These plots show how the value of a feature impacts *XplainScar* output.

*XplainScar* utilizes several ECG features for a single prediction, with only the most frequently used ones presented in **Table 4**. Top ECG features frequently selected for basal LV scar include deeper or more frequent Q waves, prolonged non-terminal QRS duration, less negative area under the QRS complex curve and smaller T wave amplitude in leads V1-V2. For apical LV scar, features such as T wave inversion, positive QRS complex area, and slope variations of the TP segment are prominent.

### LV scar burden influences *XplainScar* performance and selected features

We perform a stratified analysis to test our method’s performance based on the percentage of scar tissue mass in the LV (**Figure 2**). *XplainScar* achieves near-perfect detection in patients with scar percentage exceeding 10%. However, as scar burden decreases, there is a proportional increase in missed detections.

**Figure 2.**
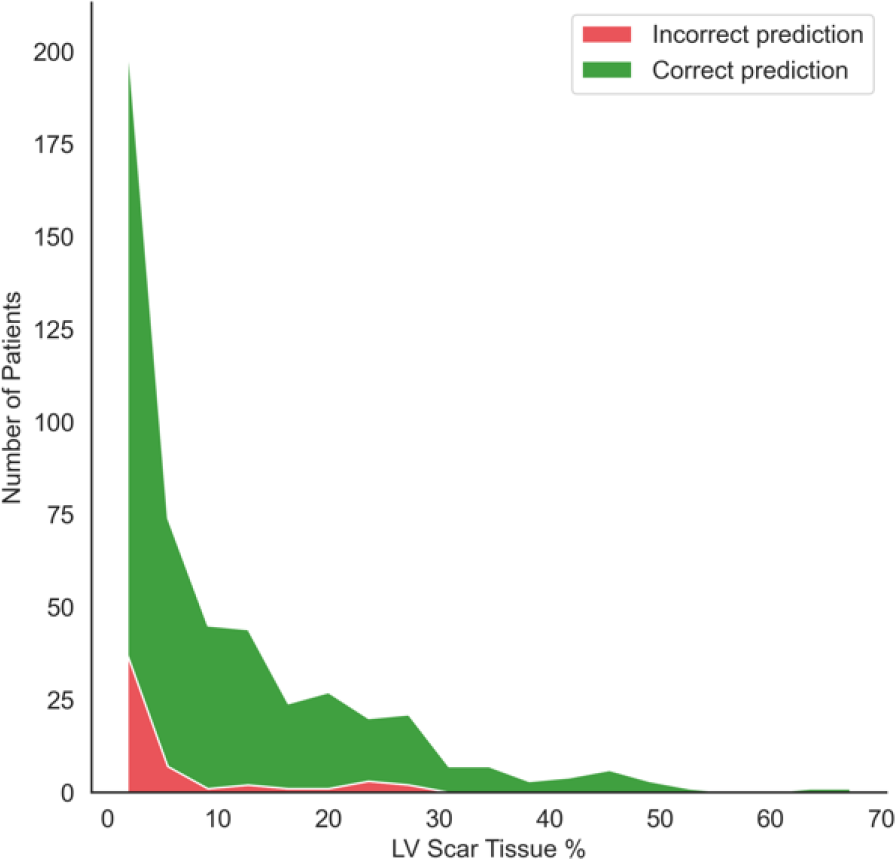
Performance of XplainScar over the JH dataset stratified by LV scar burden (scar mass as percentage of LV mass) quantified by MRI.

LV scar burden also influences the selection of ECG feature by *XplainScar.* **Supplemental Table 4A** compares frequently selected ECG features by *XplainScar* with LV scar <15% as well as >15% of LV mass, and **Table 4B** compares the value of these features between the two groups. We chose the threshold of 15% based on previous findings that demonstrated increase in sudden cardiac death risk with LV scar > 15% of LV mass.^37^ As scar burden increases, we observe a higher frequency of T wave inversion across multiple leads; Q amplitude was selected 14% more frequently in patients with LV scar burden >15%, and QRS non-terminal duration was chosen 10% of the time in patients with LV scar burden <15%.

### Case Studies

*XplainScar* is tailored to explain a single prediction. That is, given an ECG, *XplainScar* predicts the presence of scar in basal, mid, and apical regions of the LV and explains why such a prediction is made (**Figure 3**). Column-b of **Figure 3** illustrates the automatic annotation, and segmentation of the QRS complex, ST segment, T wave, TP segment, whereas column-c summarizes the ECG features with the top 20% of Shapley values, that drive scar prediction. In ECG #1, *XplainScar* used R upstroke slope in aVR, absence of S wave (*S*_*amplitude*_ = 0 *mV*), deep S wave in lead V3, fragmentation of QRS complex in V1 to predict basal-LV scar. In ECG #2 and ECG #3, scar prediction in the mid and apical LV, respectively, was driven by T wave features. T wave inversion in V2 – V6 was prominent in apical scar prediction, and T wave amplitude in V5, V6, aVL, aVR for scar detection in mid-LV.

**Figure 3.**
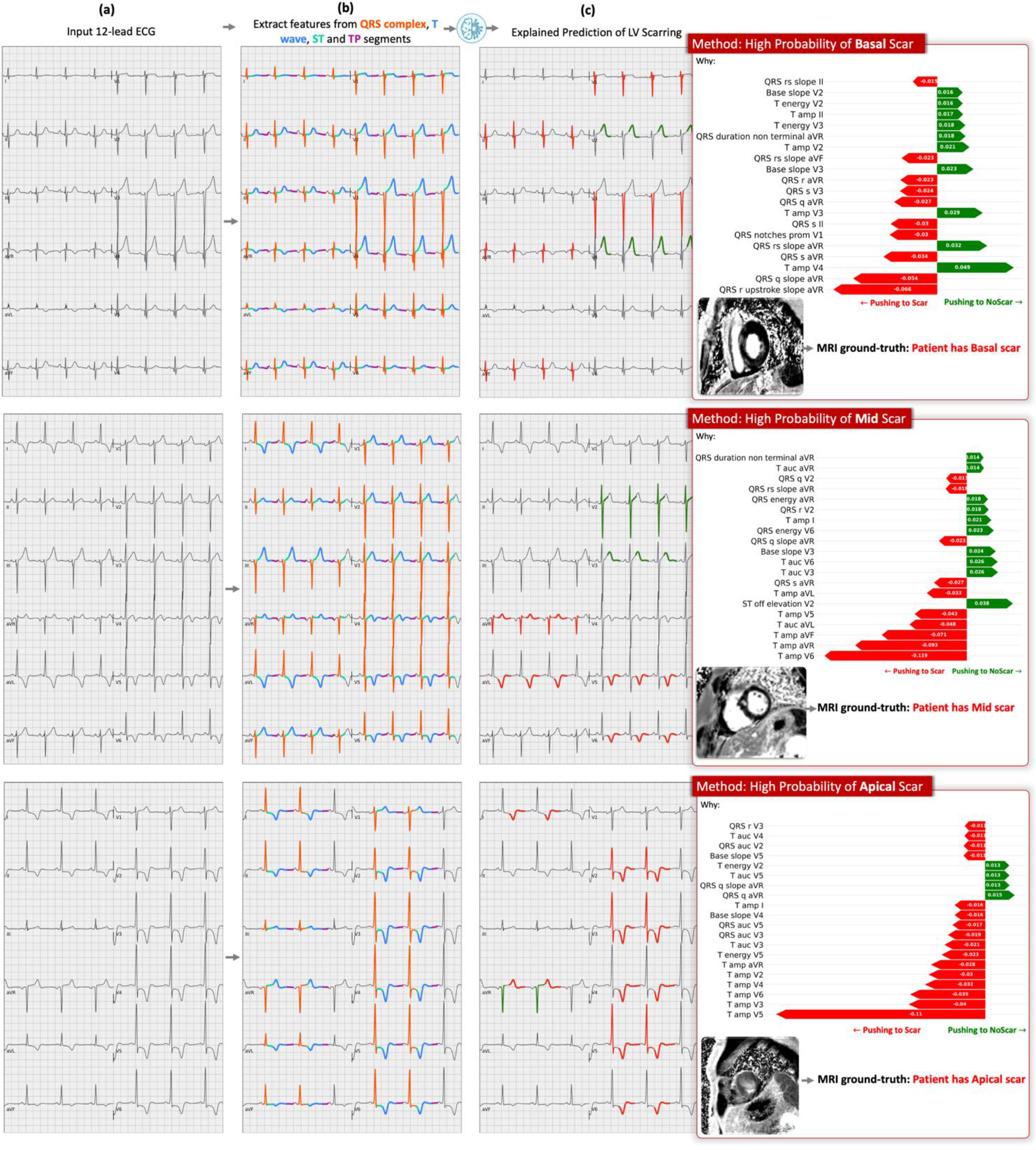
Case studies. Explained prediction of LV scar in 3 representative ECGs from the UCSF dataset by *XplainScar*. (a) Input 12-lead ECG, (b) Segmented ECG: QRS complex (in orange), ST segment (in green), T-wave (in blue), TP segment (in purple), (c) Our method’s prediction along with the ground truth label and the top 20% Shapley values showing the importance of each ECG feature in the prediction. The ECG component pushing the method towards predicting scar is highlighted in red on the 12-lead ECG, whereas those pushing the method toward outputting NoScar are highlighted in green.

### Correlation with EHR, ECHO, MRI data

We explored associations between patient clusters identified using ECG features and clinical/imaging features. To achieve this, we trained fully connected neural networks using 32 HCM variables (**Suppl. Table 5**) drawn from electronic health records (EHR), echocardiography (ECHO), and MRI data available for all patients in the JH dataset. These features are inputted into three fully connected neural networks trained on the JH dataset, learning a binary classification to identify patient cluster labels assigned during ECG partitioning (**Figure 4**) for each basal, mid, and apical LV scar detection tasks. Each network performs binary classification mapping EHR, ECHO, MRI parameters to cluster labels, evaluated using a 10-run 5-fold cross-validation scheme, with two clusters identified for basal, mid, and apical LV scar detection tasks.

**Figure 4.**
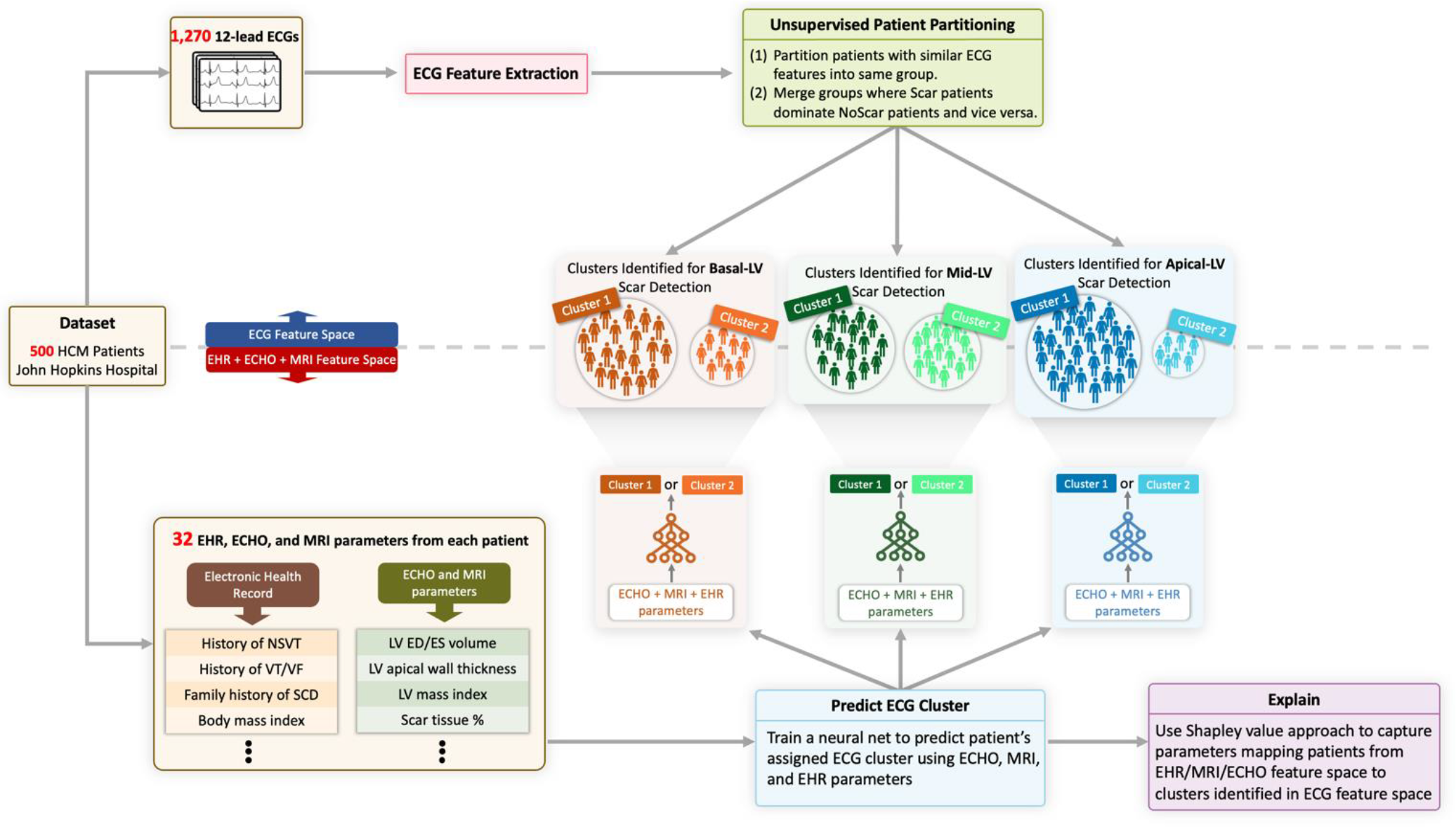
Clusters of HCM patients identified using ECG clustering correlate with EHR, ECHO, MRI data. Supervised neural networks trained in EHR, ECHO, and MRI feature space can effectively separate HCM patients into the same groups obtained by unsupervised patient partitioning step using ECG data.

Following a 10-run 5-fold cross-validation scheme, we observe that the neural networks perform well, with an average F1 score of 0.86 ± 0.03. This result suggests that patient clusters, initially identified solely through ECG analysis, can also be effectively separated using EHR, ECHO and MRI parameters, with the trained neural networks providing insights into this separation. The results presented in **Suppl. Table 6** highlight the top 5 clinical parameters selected by the neural networks to predict the cluster labels. The Shapley value of each parameter and their value (mean and standard deviation) in each cluster are shown. Notably, univariate analysis does not reveal any statistically significant differences in the values of clinical parameters between the clusters. However, the neural networks successfully capture the correlations among several clinical parameters, enabling the mapping of patients from the EHR, ECHO and MRI feature space to their respective cluster labels found in the ECG feature space.

### Longitudinal Testing

Since LGE represents cardiac replacement fibrosis, which is irreversible, we hypothesized that *XplainScar* would continue to detect LV scar in the same location, using ECGs obtained several years after initial ECG/MRI. To test this hypothesis, we identified 124 patients in the JH dataset who had ECGs recorded ≥ 4 years after MRI, with 86/124 having LV scar based on MRI. *XplainScar* detected LV scar in 74/86 patients with LV scar detected by MRI, and 13/35 patients who had no evidence of LV scar by MRI, at the time of their first clinic visit. However, none of these 13 patients had follow up MRIs, so we are unable to confirm that they developed LV scar over their follow up period. Prospective testing of our method with MRI is needed to demonstrate the utility of *XplainScar* for longitudinal monitoring of LV scar in HCM.

## Discussion

We developed a machine learning framework, *XplainScar* that is capable of handling diverse ECG patterns within a large HCM patient cohort, for effective LV scar detection and explanation. We devise two novel strategies, namely, unbiased ECG feature extraction and ECG representation-learning to equip *XplainScar* with the ability to handle highly heterogenous HCM-ECG patterns and detect scar. The first strategy we employ is to extract basic yet comprehensive ECG features rather than *compound* ones such as the Selvester QRS score [39] and fragmented QRS complex.^38^ Such compound features are calculated based on a set of basic, simple ECG criteria; however, this calculation is often challenging even for human experts, and precludes discovery of new ECG features of scar. For instance, Vandenberk et al. reported significant interobserver variability (Kappa of 0.65) among five experienced observers when identifying fragmented QRS in 100 ECGs.^39^ Compound ECG features can also introduce bias to the input feature space causing machine learning to learn less from other features that might be informative but not captured by the compound features. *XplainScar* uses a wide range of basic ECG features, based on physiological, biophysical and mathematical principles, to predict LV scar. By avoiding compound features and utilizing easily interpretable metrics, such as wave energy and amplitude, our method captures important markers for LV scar detection. Notably, our analysis agrees with previous studies regarding the significance of T wave inversion in apical LV scar.^40,41^ Our method also uncovers previously unexplored ECG markers of LV scar, such as T wave energy, area under QRS, and Q amplitude, which play a significant role in detecting basal scar.

Our second strategy to handle highly heterogenous HCM-ECG patterns is the novel combination of unsupervised and self-supervised representation learning. Unsupervised ECG clustering has been successfully employed to overcome challenges facing the ECG supervised learning classification, for instance, to resolve the imbalanced data problem and low-level automation of patient-specific ECG classifiers.^42^ Motivated by these applications, we propose a novel yet simple unsupervised approach to reduce inter-patient variability among HCM patients while increasing separation between the Scar and NoScar groups of patients. Our method partitions patients into groups consisting of several sub-clusters each containing patients with similar ECG patterns (low inter-patient variability), where the number of patients belonging to the Scar or NoScar class dominate the other class (high separation between classes). Although we identify groups of patients with high separation between the Scar and NoScar classes, an informative ECG representation discriminating the two classes is still needed. Such representation is challenging to learn as the number of training samples and scar labels to train a performant classifier is limited in each group. Our self-supervised representation learning addresses this challenge. Self-supervised learning has proved effective in classifying raw ECG signals into abnormality classes despite scarce expert-provided labels.^43–45^ Here, we employ it to learn a useful representation from tabular ECG features in each group, facilitating downstream scar classification. We couple self-supervised learning with a data augmentation algorithm (SMOTE, explained in the Methods section) to effectively increase the size of the training dataset. The augmentation algorithm proved to be highly effective for self-supervised learning. This can be due to the low inter-patient variability achieved by our unsupervised learning approach: the datapoint synthesized by interpolating two real datapoints from class *c* is more likely to belong to *c* due to the low inter-patient variability, and consequently less likely to disturb the governing class distributions.

Our ablation experiments clearly demonstrate that the combination of unsupervised ECG clustering and self-supervised representation learning is crucial for effective LV scar detection. Importantly, the groups of patients identified by unsupervised ECG clustering are not arbitrary partitions but exhibit shared clinical parameters, even though these parameters were not seen during clustering. Notably, a supervised neural network trained on patients’ EHR, ECHO and MRI parameters successfully segregates patients into the same groups identified by ECG clustering. This mapping of patients from one modality to another provides an extra layer of validation for *XplainScar*. The incorporation of clinical parameters and the correlations captured by the neural networks can pave the way for enhanced LV scar detection and personalized patient management. By integrating information from multiple modalities, *XplainScar* facilitates a deeper understanding of the complex relationships between ECG patterns, clinical parameters, and LV scar. The combination of unsupervised ECG clustering and self-supervised representation learning and bridging between multiple modalities are novel approaches that have not been investigated before in other applications of computational ECG analysis. Thus, our work opens new avenues for future research in the field.

A key strength of *XplainScar* lies in its explanation framework, which provides ECG signatures of basal, mid and apical LV scar in HCM, and thus prompts user trust in our method. Our explanation framework also sheds light on the inherent combinatorial non-linear power of *XplainScar*. Through this framework, we have discovered important ECG features of LV scar that do not exhibit a significant difference between the Scar and NoScar groups but are frequently selected by *XplainScar*. For example, non-terminal QRS duration (also called intrinsicoid deflection) is frequently selected by our method when predicting basal-LV scar, but does not exhibit a significant difference between Scar and NoScar groups (*p* = 0.12).^46^ We use SHAP to explain our model’s predictions of LV scar; SHAP employs a linear function that assigns importance scores to individual features but does not elucidate the relationships among these features.

An important translational application of *XplainScar* is the ability of our model to uncover ECG signatures of LV scar in each HCM patient. To the best of our knowledge, there is no existing method that extracts comprehensive ECG features representing ventricular depolarization and repolarization from all 12 leads of a routine rest ECG, and provides a signature of scar in the basal, mid and apical LV in HCM. Prior studies have mainly focused on abnormalities in the QRS complex and/or T waves to derive scores that predict LV scar in heart disease.^40,47–50^ However, given the heterogeneity in the location and degree of cardiac hypertrophy, fibrosis as well as the diversity of ECG abnormalities in HCM patients, machine learning methods that use all 12 leads of the ECG are best suited for LV scar detection and explanation.

Each lead of the 12-lead ECG records the electrical field generated by myocardial depolarization and repolarization in a specific region of the heart: V1-V2 reflect electrical activity in the septum; V2- V4 reflect anterior wall activity; I, aVL, V5-V6 reflect lateral wall activity; aVR reflects activity in the basal septum and RV outflow tract. Hence, abnormalities in specific leads are useful to localize cardiac pathology such as fibrosis (scar) which can slow impulse propagation and promote dispersion of repolarization.^51,52^ Higher scar burden would be expected to have greater impact on cardiac electrophysiology, which could explain our results of model performance association with LV scar percentage (**Figure 2**). An interesting and novel result of our study is the identification of several QRS features in lead aVR for scar prediction in the basal LV (**Figure 3**, **Table 3**); these results are physiologically accurate but would likely have been missed if manual ECG measurements had been performed, because lead aVR is often ignored by clinicians during ECG analysis.^53,54^ Apical LV scar prediction by *XplainScar* was dominated by T wave features in leads V2-V6 (**Figure 3**, **Table 3**), which has been confirmed by prior ECG-based studies.^40,41^ While T wave inversions in several leads are common in HCM, deep T wave inversions are most commonly seen in apical HCM. Our results from *XplainScar* indicate that fibrosis likely plays a role in generation of deep T wave inversion in leads (V4- V6) that reflect electrical activity in the cardiac apex, by causing inversion of the repolarization sequence.^52^ Thus, *XplainScar* has the potential to assist clinicians with assessing and monitoring development of electrical and structural LV remodeling in HCM.

## Limitations

This is a retrospective study, using multi-center ECG data for detection of LV scar at the time of patients’ first clinic visit. Prospective multicenter studies combining ECG and MRI are needed to explore the potential of *XplainScar* for longitudinal scar detection in HCM. While ECG features identified by *XplainScar* vary with LV scar burden, it is not designed to quantify LV scar. Future model development for longitudinal monitoring and quantification of LV scar would be useful to assist with risk stratification for sudden cardiac death, disease management and testing of antifibrotic therapies.

## Conclusions

*XplainScar,* the first explainable ECG-based machine learning method for LV scar detection and localization in HCM, is computationally lightweight and demonstrates high performance with an F1 score > 90%. *XplainScar* identified ECG signatures of scar in the basal, mid, and apical LV, which included a large number of new physiologically accurate ECG features.

## Data Availability

XplainScar is publicly available at https://github.com/KasraNezamabadi/XplainScar

## Acknowledgments

We would like to express our gratitude to Dr. Hagit Shatkay, Dr. Geoff Tison, Dr. Jeffrey Olgin for their guidance, support and expertise.

